# Modelling the impact of hybrid immunity on future COVID-19 epidemic waves

**DOI:** 10.1101/2023.03.12.23287174

**Authors:** Thao P. Le, Isobel Abell, Eamon Conway, Patricia T. Campbell, Alexandra B. Hogan, Michael J. Lydeamore, Jodie McVernon, Ivo Mueller, Camelia R. Walker, Christopher M. Baker

## Abstract

Since the emergence of SARS-CoV-2 (COVID-19), there have been multiple waves of infection and multiple rounds of vaccination rollouts. Both prior infection and vaccination can prevent future infection and reduce severity of outcomes, combining to form hybrid immunity against COVID-19 at the individual and population level. Here, we explore how different combinations of hybrid immunity affect the size and severity of near-future Omicron waves. To investigate the role of hybrid immunity, we use an agent-based model of COVID-19 transmission with waning immunity to simulate outbreaks in populations with varied past attack rates and past vaccine coverages, basing the demographics and past histories on the World Health Organization (WHO) Western Pacific Region (WPR). We find that if the past infection immunity is high but vaccination levels are low, then the secondary outbreak with the same variant can occur within a few months after the first outbreak; meanwhile, high vaccination levels can suppress near-term outbreaks and delay the second wave. Additionally, hybrid immunity has limited impact on future COVID-19 waves with immune-escape variants. Enhanced understanding of the interplay between infection and vaccine exposure can aid anticipation of future epidemic activity due to current and emergent variants, including the likely impact of responsive vaccine interventions.

## 1 Introduction

The global spread of SARS-CoV-2, causing COVID-19 disease, has fundamentally changed society. Multiple epidemic waves have been experienced, beginning with the wild-type virus in early 2020, and followed by the emergence of variants such as Alpha (B.1.1.7), Delta (B.1.617.2), and more recently Omicron (B.1.1.529, subvariants BA.1, BA.2, BA.3, BA.4, BA.5 and descendent lineages) [1]. In the absence of specific preventive or disease modifying agents, the only initially effective measures to reduce the impact and burden of COVID-19 were case and contact management, restrictions to limit the number of social interactions and personal protective behaviours to reduce the per-contact likelihood of transmission. The initial hope was that vaccines might provide long lasting immunity against infection, with definitive impacts on transmission. However, while efficacy against severe disease appears to be relatively robust and long lasting, efficacy against infection acquisition and onwards transmission are lower and shorter lived—especially against the Omicron variant [2]. In response to repeated epidemic waves in highly immune populations, one or more booster doses have been recommended for sustained protection [3]. Omicron and its subvariants have caused high levels of exposure in multiple populations world-wide, even in highly vaccinated populations [4], and has led to complex “immune landscapes” across the world with varying levels of so-called “hybrid immunity” [5] in vaccinated and infected populations, consisting of immunity derived from both past infection and past vaccination.

Both infection and vaccination provide immunity boosting effects to individuals [6, 7, 8, 9, 10, 11, 12, 13] and indirect benefits to others in the population, regardless of vaccination status, through reduction in onwards transmission [14]. This immunity directly reduces the risk of infection, and, if infection does occur, reduces disease severity and lowers onward transmission. Neutralising antibody titres, which are boosted by exposure to infection and/or vaccination, are correlated with efficacy against clinical endpoints of infection, symptoms and severe disease outcomes [15, 16, 17]. However, infection and vaccination can give different levels of protection [18], which are variable across demographics (especially with respect to age) [19]. Increased breadth and duration of antibody responses have been observed in individuals who have been both infected and vaccinated, termed “hybrid immunity” [20, 21, 22, 23, 24, 25]. Crucially though, all forms of immunity *decay over time*, with additional complexity arising from observations of differential antibody waning following infection, vaccination or a combination of the two [26, 27]. Furthermore, the recent Omicron subvariants have shown immune escape in relation to immunity derived from past infections and vaccinations [28, 29, 2, 30, 31], and even infection from earlier Omicron subvariants (BA.1, BA.2) has been shown to have reduced protection against later Omicron subvariants (BA.4, BA.5) [32].

Overall, populations have hybrid immunity containing multiple groups of individuals with different vaccination and past-infection statuses. It is a challenging process to synthesise information about individual-level immunity to population-level protection and make predictions about the severity of future COVID-19 waves and optimisation of primary and booster vaccine allocation [33, 34, 35, 36, 37]. Modelling has been used to support decision making around the world in regards to managing COVID-19 [38, 39, 40]. It has been used extensively to compare different vaccination strategies [41, 42, 43], but many either do not include waning of immunity [44], or do not take a hybrid-immunity approach [45]. However, the inclusion of both of these factors is key in understanding the combined population-level effect of vaccination and prior exposure upon future transmission dynamics [34, 46]. Furthermore, two populations with apparently similar levels of past-infection and vaccination coverage could still have different responses, as *epidemic history*, including the SARS-CoV-2 strain(s) and exposure sequence, also play an important role [47]. Hence, different populations and regions with unique epidemic histories require individual analysis.

In this study, we focus on dynamics of immunity in the World Health Organization (WHO) Western Pacific Region (WPR). Prior to the rollout of vaccination, the WPR region had low seroprevalence compared to other regions such as Europe [48]. As such, infection-derived immunity within this region is now largely a result of the recent Omicron wave, with relatively few infections caused by previous variant strains, unlike the populations with high prior Delta exposure pre-Omicron [34, 44]. Hence, our focus will be on analysing scenarios where a population’s past infection-derived immunity is due to Omicron.

We use an agent-based (individual-based) model of COVID-19 to consider how combinations of vaccination coverage and prior Omicron infection exposure can protect a population from future near-term Omicron waves, in the context of fixed public health measures (or lack thereof), no TTIQ (test, trace, isolate, quarantine), and with a constant vaccination capacity. We focus on the interplay between younger and older population demographics—as a proxy for two populations with different high-risk group sizes—along with a range of vaccination coverages and prior infection rates. Understanding hybrid immunity is necessary to develop efficient allocation of vaccine resources to achieve equity of outcomes across different populations with unique hybrid immunity statuses, and will also allow us to understand larger-scale future societal impacts, such as worker absenteeism and overloaded health systems. Given that past infection and vaccination provide unequal levels of immunity, we aim to identify the most advantageous strategies to protect populations from infection and severe disease outcomes in future, taking into account the current population immunity profile.

## 2 Methods

The overall simulation procedure is comprised of an infection transmission/dynamics model that is linked to a mechanical agent-based model [49, 50]. Outputs from this model feed into a clinical pathways model. The simulation process is depicted in Fig 1, with individual model components described in more detail below.

**Figure 1:**
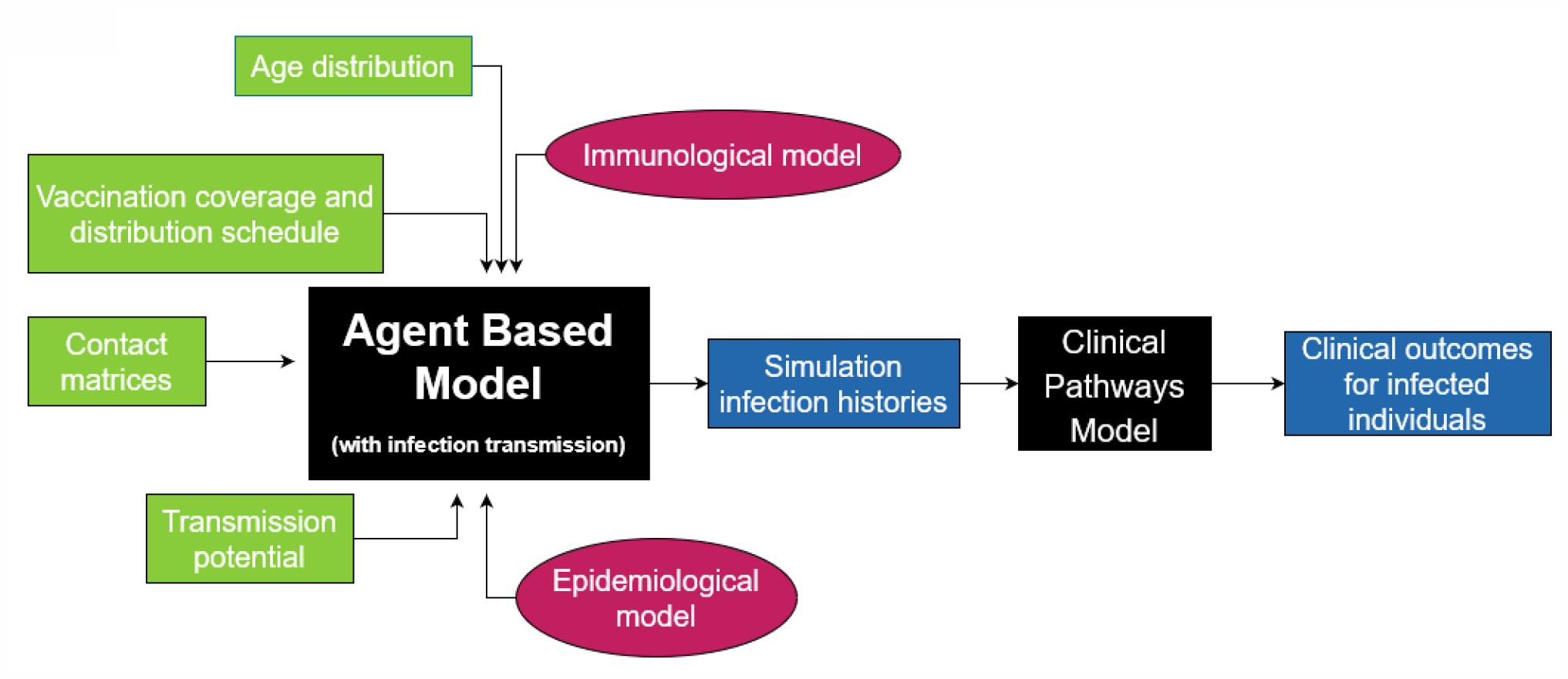
Diagram of overall simulation procedure. The core of the simulation uses an agent-based model with an underlying infection transmission model, with multiple primary inputs including immunological parameters and scenario-demography setups. The outputs are then fed into a clinical pathways model that produces clinical outcomes for infected individuals.

### Agent-based transmission model

We use the agent-based COVID-19 model of Conway et al. [49, 50]. In the agent-based model, each individual has a specific age, unique neutralising antibody titre, vaccination history and own compartment label susceptible/exposed/infected. At the beginning of each simulation, all individuals have zero neutralising antibody titre. An individual’s neutralising antibody titre can be boosted due to infection and/or vaccination and decay over time, with different levels of boosting depending on vaccine product and infection variant.

In the model, infection transmission occurs due to contact between infectious and susceptible individuals, which is dependent on input contact matrices and the individuals’ ages. Once infected, the model randomly samples to determine whether an individual is asymptomatic or symptomatic, as well as generating when they become infectious, time of symptom onset, time of isolation (if isolation or quarantine is included), and time of recovery, based on parameter values in Table 1. In each simulation, this information can be exported as a line list of infections.

**Table 1:** Model parameters. Note that the age brackets for probabilities of symptomatic infection (prob_symptoms), relative infectiousness once infectious (relative_infectiousness), and susceptibility to becoming infected upon contact with an infected individual correspond to [0, 5, 10, 15, 20, 25, 30, 35,40, 45, 50, 55, 60, 65, 70, 75, 80]. “*c*_50_” corresponds to the midpoint of the logistic function for a particular disease outcome (hospitalisation, death etc…). Source: Refs. [49, 50, 53].

| Parameter: description | Value |
| --- | --- |
| $c_h$: $c_{50}$ of hospitalisation | -1.22 |
| $c_d$: $c_{50}$ of death | -1.18 |
| $c_\xi$: $c_{50}$ of acquisition | -0.472 |
| $c_\tau$: $c_{50}$ of transmission | 0.0295 |
| $c_q$: $c_{50}$ of symptoms | -0.644 |
| $\mu_{AZ1}^0$: $\log_{10}$ of the mean neutralising antibody titre after the first dose of AstraZeneca (no infection) | -0.530 |
| $\mu_{AZ2}^0$: $\log_{10}$ of the mean neutralising antibody titre after the second dose of AstraZeneca (no infection) | -0.120 |
| $\mu_{PB2}^0$: $\log_{10}$ of the mean neutralising antibody titre after the second dose of Pfizer (no infection); also the mean titre after one AZ dose and one infection | 0.154 |
| $\mu_B^0$: $\log_{10}$ of the mean neutralising antibody titre after the first mRNA booster dose (both with and without infection); also the mean titre after two AZ doses and one infection | 0.323 |
| $\mu_U^0$: $\log_{10}$ of the mean neutralising antibody titre after infection whilst unvaccinated | 0 |
| $\log(k)$: governs the logistic curve steepness relating antibodies to protection against disease outcome | 1.69 |
| $k_a$: decay rate of neutralising antibodies | 0.00824 |
| $\log_{10}(f_{\text{Omicron}})$: $\log_{10}$ of the fold change in neutralising antibody titre between Delta and Omicron (BA1- like) | -0.692 |
| $\sigma$: standard deviation of the $\log_{10}$ of neutralising antibodies across the population | 0.465 |
| probability of symptoms across age groups | [0.29, 0.29, 0.21, 0.21, 0.27, 0.27, 0.33, 0.33, 0.4, 0.4, 0.49, 0.49, 0.63, 0.63, 0.69, 0.69, 0.69] |
| relative infectiousness across age groups | [0.799, 0.688, 0.675, 0.756, 0.918, 0.965, 0.947, 0.932, 0.934, 0.940, 0.954, 0.982, 1.0, 0.998, 0.990, 0.974, 0.944] |
| susceptibility across age groups | [0.301, 0.367, 0.433, 0.527, 0.764, 0.924, 0.983, 0.974, 0.932, 0.915, 0.929, 0.962, 1.0, 0.972, 0.882, 0.824, 0.802] |
| $\log_{10}(f_{\text{Omicron-escape}})$: $\log_{10}$ of the fold change in neutralising antibody titre between Delta and the BA4/5-like immune escape variant | -1.18 |
| $R_0$ ratio between the original Omicron variant (BA1/2-like) and the BA4/5-like immune escape variant | 1.3 |

### Clinical pathways model

We use the stochastic COVID-19 clinical pathways model by Conway et al. [49]. The clinical pathways model relates demographic information and neutralising antibody titre to clinical outcomes. For each symptomatic infection occurrence, the clinical pathways model takes in age and neutralising antibody titre at time of infection of the infected person and generates a clinical trajectory for each symptomatic infection, including whether the individual will require hospital admission, or will die.

Note that the clinical pathways model is independent of the main agent-based model described above, which allows for greater flexibility in the overall simulation process and also allows the clinical pathways model to be used for multiple epidemic models. However, due to this independence, it is possible that patients who die, according to the clinical pathways model, remain in the agent-based simulation. Due to the low number of deaths, relative to infections, this should not have major effect on the two models’ outcomes [49].

### Immunological model

Neutralising antibodies are one of the many biomarkers associated with COVID-19 immunity [51, 18, 52], so we use them to regulate each agent/individual’s interaction with the SARS-CoV-2 virus, including each individual’s level of protection against infection and severe clinical outcomes [53, 15, 16]. In particular, we use the model from [15, 16] to use neutralising antibodies to determine each individual’s level of protection against infection, symptomatic disease, onward transmission given breakthrough infection, hospitalisation and ICU admission, and death. The antibodies follow exponential decay over time, with our model assuming that the decay rate is the same across all forms of initial antibody boosting (whether vaccination or prior infection).

When an antibody boosting event occurs, the individual gets a new antibody titre, *a*^0^, that is sampled from:

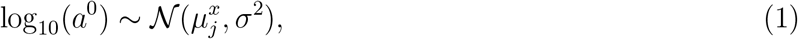

where 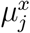 is the mean neutralising antibody titre against strain *x* (Delta or Omicron) after boosting process *j* (infection or vaccination), and *σ*^2^ is the variance of antibodies across the population (Table 1). Note that infection prior or post vaccination results in the same titre. The neutralising antibody titre boosting occurs with the booster dose, and does not rise with extra infections. There is an upper limit on titre, which is equivalent to either two vaccinations and an infection, or three vaccinations. (For further details on how the titres for determined for combinations of infection and vaccination, see Table 1 of [50].)

Note that there is an additional scaling as many of the parameters in Table 1 are for strain 0, i.e. the Delta strain. To convert to neutralising antibodies against the Omicron strain, the fold change *f*_Omicron_ is used:

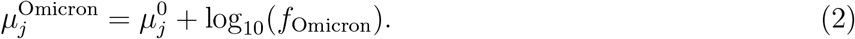

To convert an individuals’ Omicron BA.1 titres, *a*^*Omicron*^, to titres against the BA.4/5-like variant:

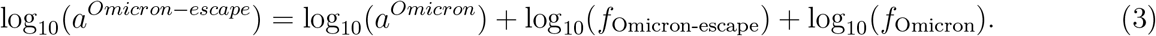

The immunological model parameters are drawn from Refs. [49, 53, 50] (Table 1). The model parameters broadly follow the characteristics for the Omicron BA.1 subtype. We include two additional parameters in the case of an Omicron BA.4/BA.5-like immune escape variant: 1) *R*_0_ ratio between the original Omicron variant (BA1/2-like) and the BA4/BA5-like immune escape variant to account for an inherent increase in transmissibility; and 2) log_10_ (*f*_Omicron-escape_) to account for an approximately three-fold reduction in neutralising antibody protection derived from preceding immunising exposures [54, 53].

The relationship between neutralising antibody titre *a*_*i*_ of an individual to protection against some disease outcome *ρ*_*α*_ (hospitalisation, death, acquisition etc.) is [49, 50]:

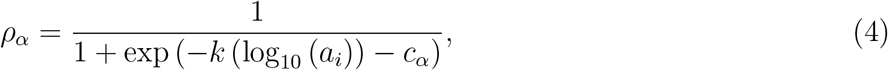

where *k* determines the steepness and *c*_*α*_ corresponds to the midpoint of the logistic function for a particular disease outcome (*α* = hospitalisation, death etc…) (see Table 1).

### Fixed input parameters

In the WPR scenarios we consider, the following components are fixed:

#### Vaccine type

We assume that all primary doses are ChAdOx1 nCoV-19 (AstraZeneca) and booster doses are BNT162b2 (Pfizer/BioNTech).

No **TTIQ** (Test, Trace, Isolate and Quarantine). This means there are no public health measures in these scenarios.

#### Timeline

We assume a fixed schedule for vaccination and the seeding of infection (depicted in Fig 2).

**Figure 2:**
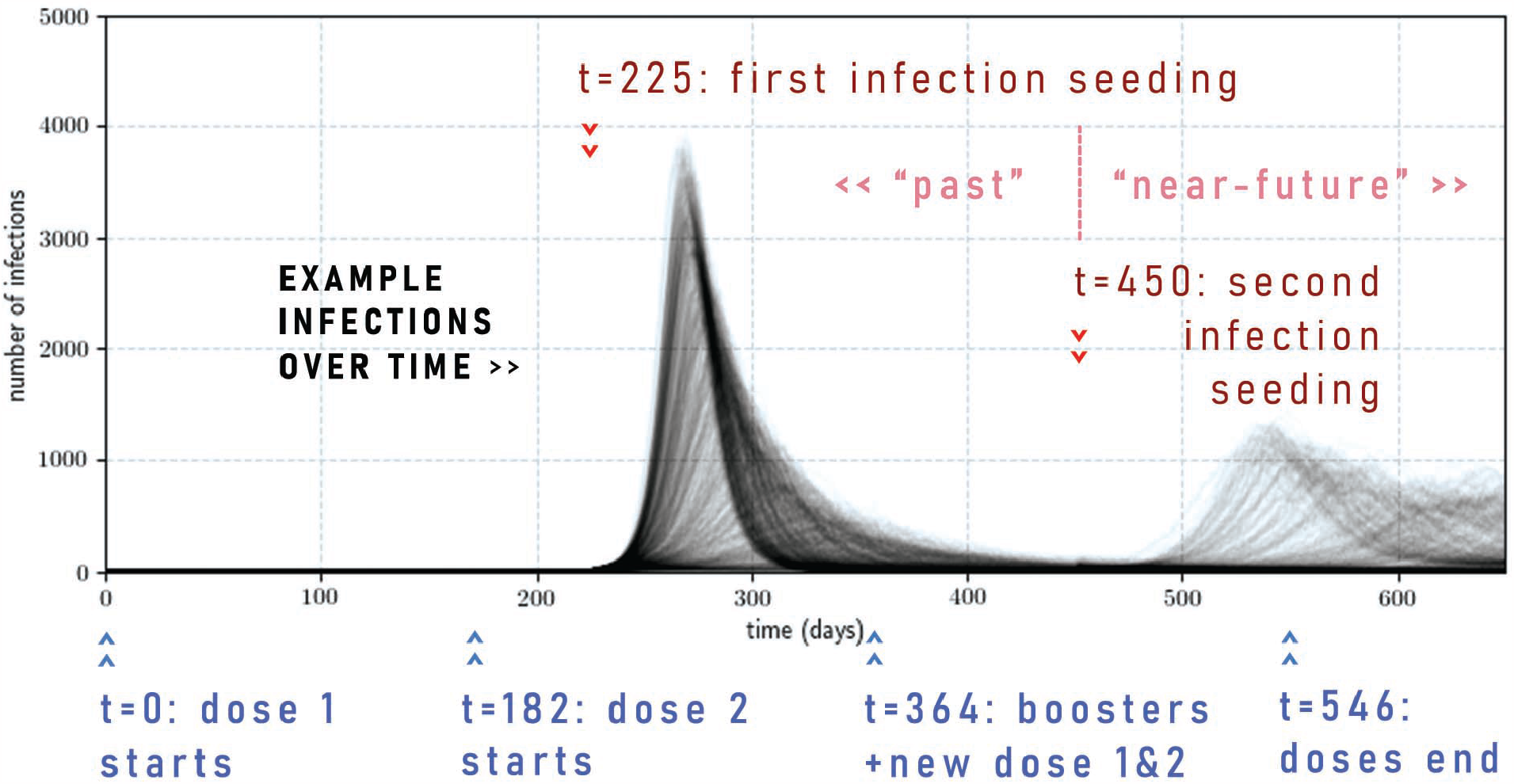
Timeline of vaccination schedule and infection seedings with examples of infection time series. There is a vaccination rollout that occurs in three consecutive stages, starting at *t* = 0, *t* = 182, and *t* = 364 and ending at *t* = 546. The first wave and the second wave are generated by randomly seeding 100 infections in the population (which could occur due to a super-spreader event, for example) at times *t* = 225 and *t* = 450. The example time series are for a second wave due to the same variant as the first wave.

There are three vaccination stages that take 26 weeks each: the first dose stage, the second dose stage, and the mixed booster and new primary course stage. In the first stage, first doses are given to the allocated population, and in the second stage, the second dose is given to the same groups. Within the first and second stages, the 65+ age group always gets their first dose or second dose first, followed by randomly assigned vaccinations among the 5 *−* 64 age group. At the end of the second stage, *V* % of the population have received the primary course, where *V* = 20, 50, 80. In the third stage, 80% of the fully vaccinated population receive a booster dose, while remaining available doses are given out as new primary doses. An example of this is given in Fig. 9.

We seed two infection waves with 100 infections each: one at time *t* = 225 (approximately 32nd week), such that the first wave is largely over by the end of the second vaccination stage, and one at time *t* = 450, where protection from the first wave has waned enough such that a second wave could possibly occur. Any infection that occurs before *t* = 450 is marked as a “past” infection, while anything after *t* = 450 (and before *t* = 650) is marked as a “near-future” infection.

Note that we end our analysis at time *t* = 650, which is approximately three months after the final vaccination stage finishes. We do not run simulations for any extra time until all second waves are finished in situations where the second seeding does not spark a second wave immediately. This is because if we do continue to run the simulation, immunity will continue to wane, and in some situations, we would see a third wave. Furthermore, we would expect that some populations would continue a fourth vaccination stage and so forth.

### Scenarios

The scenarios and demographics that we change across different simulations are as follows.

#### Population age demographics

see Fig. 3(a) and (b). We generate populations with 100,000 people. We consider “older” and “younger” populations, which are derived from averaged age-proportions across the majority of countries in the Western Pacific regions, using the population data acquired through https://population.un.org/wpp/DataQuery/ [55] for the year 2021. We defined “older” countries as those with an *OADR* ≥ 15 and “younger” countries as having an *OADR* ≤ 12, where OADR is the *old-age to working-age demographic ratio*, which we calculate as a ratio between the +65 year old population and the 20-64 year old population. The individual country-level data used can be seen in Fig. 6 and Fig. 7.

**Figure 3:**
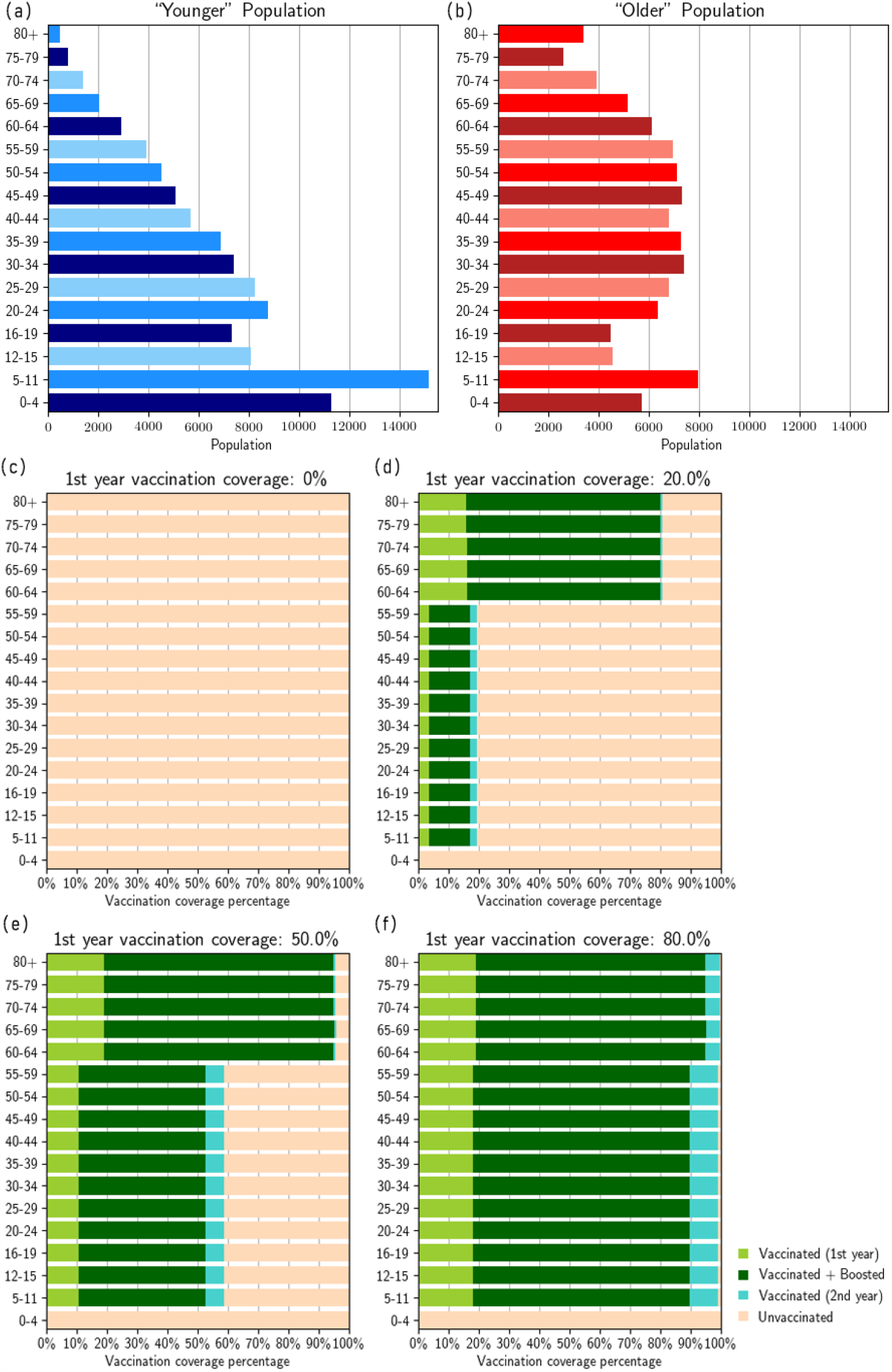
Scenarios considered: (a) exemplar ‘younger’ population demographics, (b) ‘older’ population demographics (see Appendix A for the construction of these exemplar populations), (c) no vaccination, (d) 20% vaccination coverage, (e) 50% vaccination coverage, (f) 80% vaccination coverage, where the coverage value corresponds to primary vaccination coverage by time *t* = 364.

Additionally, each older/younger demographic has a different **contact matrix**, which governs the social mixing (and thus infection spread) in the model. To derive some exemplar contact matrices, we used aggregated contact matrices from “older” (*OADR* ≥ 15) and “younger” (*OADR* ≤ 12) countries that could be found on http://www.socialcontactdata.org/ [56, 57, 58, 59, 60, 61, 62, 63, 64, 65, 66] (where for “older” countries we included Belgium, Finland, France, Germany, Hong Kong, Italy, Luxembourg, Poland; and “younger” included Viet Nam and Zimbabwe. These countries fell into the appropriate OADR values and had contact matrix values for all the age-groups we required.)

#### Vaccination coverage

see Fig. 3(c), (d), (e), and (f). The vaccine allocation broadly follows WHO guidelines [3], which recommends prioritising the vaccination of older and higher-risk groups. Aside from the zero-vaccination scenario, either 20%, 50%, or 80% total vaccination coverage is achieved at the end of the second stage, i.e. *t* = 364, reflecting differing levels of vaccine access/health sector capacity. At the lower vaccination rate of 20%, we first allocate doses such that 80% of the 65+ age group are fully vaccinated by the end of the first two stages. At the higher vaccination rates of 50% and 80%, we allocate initial doses such that 95% of the 65+ age group are fully vaccinated by the end of the first two stages—a fully vaccinated cohort is unrealistic. The remaining available doses in the first two stages are then equally (proportionately) allocated to the 5 *−* 64 age groups. In the third stage, 80% of all vaccinated individuals are allocated booster doses (no prioritisation of any age groups), with the remaining doses given equally across the age-groups to unvaccinated individuals. An example of this is given in Fig. 10.

#### Second wave variant type

We considered scenarios with either a second wave caused by the same Omicron variant as the in the first wave (default scenario), or with a *new* immune escape variant. The variant in the first wave is broadly referable to the characteristics of the Omicron BA.1 variant. The immune escape variant is Omicron ‘BA.4/5-like’, as it is currently one of the variants of most global concern. The immunological parameters were determined using the methods described in [53] (also see the methods described in [49, 50]).

#### Inherent transmissibility of the variant in that particular population

Different countries do not have the same *R*_*eff*_ or *R*_0_ for the same virus. Furthermore, changes in social mixing patterns, due to social distancing measures for example, will change the population level of transmission. Therefore, rather than fixing the transmissibility across all simulations, we *vary* it (*R*_0_ ∈[0.85, 0.9, 0.95, 1., 1.05, 1.1, 1.15, 1.2, 1.25, 1.3, 1.35, 1.4, 1.45, 1.5, 1.55, 1.6, 1.65, 1.7, 1.75, 1.8, 1.85, 1.9, 1.95, 2.0, 2.05]), which gives rise to different attack rates in the population. We assume that the populations act the same during both waves (at whatever level of restrictions they may have). If the second wave is due to the same COVID-19 variant, then the transmissibility remains the same, i.e., within any individual simulation, the level of transmission is constant over time. If the second wave is due to an immune-escape variant with increased transmissibility, then the original transmissibility is multiplied up by the value given in Table 1.

For each scenario and set of input parameters, we ran 10 simulations, each producing a single infection history with information about the number and times of infection and vaccinations for each individual in the population. From each individual simulation, we produced five different clinical outcome histories with information about number of infections, severe cases, hospital admissions, ICU admissions, and deaths per day. We aggregated the total cases, ICU admissions, and deaths per day for each simulation.

## 3 Results

Fig. 4 presents results for the scenarios where both waves are due to the same Omicron BA.1-like variant. Fig. 5 presents results for the case with the immune-escape variant during the second wave. Extended figures are given in Appendix A.3.

**Figure 4:**
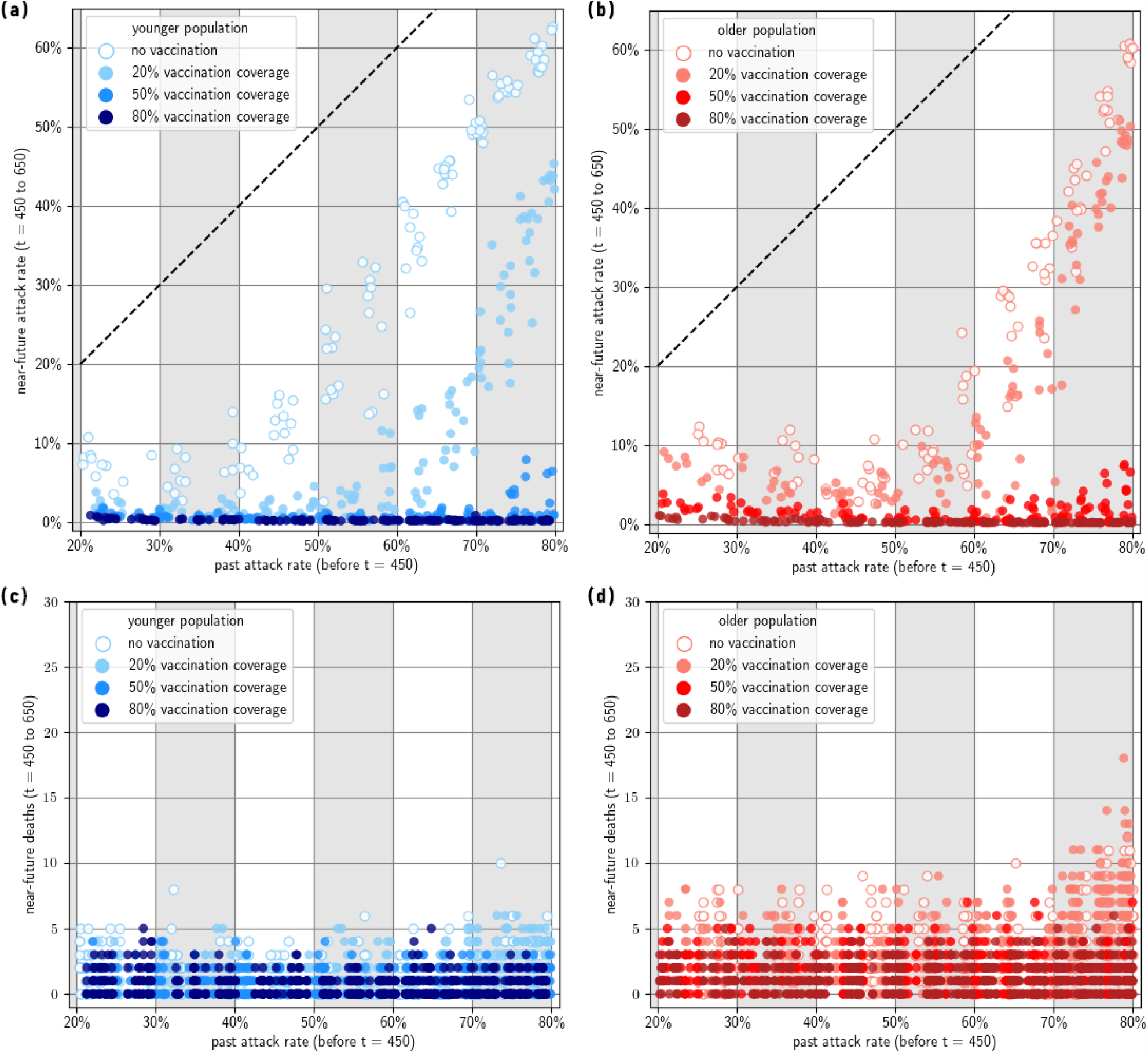
Near-future outcomes given past immunity. (a) Younger population, near-future attack rate; (b) Older population, near-future attack-rate (the diagonal line represents where past and near-future attack rates are equal); (c) Younger population, near-future deaths; (d) Older population, near-future deaths. Note that past attack rate is calculated between *t* ∈ (0, 450). Past attack rate is dependent on transmission potential, which is different for various simulations, reflecting different populations’ intrinsic transmission. Near-future attack rate and near-future deaths are calculated between *t* ∈ [450, 650]. Note that we have only included simulation results in which the past attack rate is between 20% and 80%.

**Figure 5:**
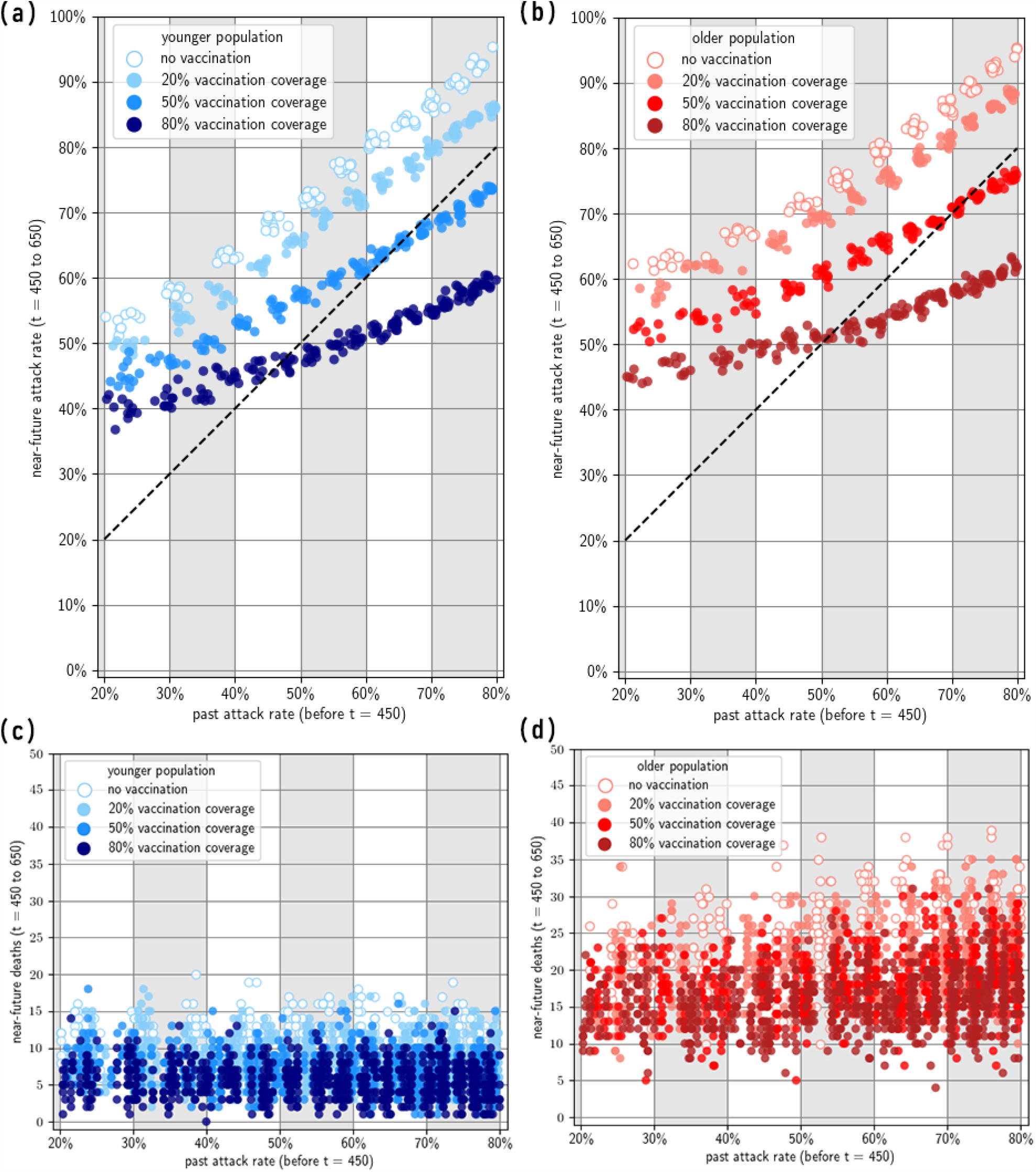
Near-future attack rate and deaths given past immunity, for a second wave due to a BA.4/BA.5-like immune escape variant. The diagonal line represents where past and near-future attack rates are equal. Note that we have only included simulation results in which the past attack rate is between 20% and 80%.

### Reinfection with the same Omicron BA.1-like variant

In the absence of vaccination, immunity from infection alone wanes rapidly enabling a second wave. Assuming that the variant characteristics are unchanged, we observe a consistent relationship linear relationship between the size of the first and second wave, especially at transmissibility values that lead to a first-wave attack rate of 50% or above. At high attack rates, the immunity derived from the first wave only gives a modest constraint on the size of the second wave (for example, a past attack rate of 80% during the first wave leads to an attack rate of 60% in the second wave) (Fig. 4 a, b).

There is little difference in the protection provided by different vaccination coverage levels when the past attack rate is 50% or lower. The past attack rate size is largely determined by the inherent transmissibility of the Omicron variant in the population. This means that for a lower past attack rate, the virus is less transmissible in that population, due to some inherent properties of the population. In countries with lower transmission, high vaccination is not as important provided that only the same variant is in circulation (Fig. 4).

For scenarios where the past attack rate is higher than 50%, we observed that different vaccination coverage levels have noticeable effects. Increasing vaccination coverage from 20% to 50% can delay the next wave in the immediate future. If vaccination coverage is only 20% then a high past attack rate is insufficient to ensure a low future resurgence of infections with the same variant. This is most likely due to the high intrinsic disease spread which caused the high past attack rate and similarly caused the high near-future attack rate, as well as the short-lived nature of immunity derived only from natural infection.

We observed little difference in total infections at vaccine coverage levels between 50% and 80%. This could be due to the fixed assumption that for each of these coverage scenarios, takeup within the 65+ age group is the same (at 95% in-group coverage). In terms of more severe clinical outcomes such as death, older populations have consistently worse clinical outcomes than the example younger population. Furthermore, a very high vaccination rate of 80% does have noticeably better clinical outcomes than 50%, especially in older populations. Since the high-risk coverage is the same in both scenarios, this indicated that there is benefit to vaccinating low-risk groups (Fig. 4 c, d).

### Reinfection with an Omicron BA4/BA5-like immune escape variant

In general, we found that for scenarios where the second wave is dominated by the Omicron BA.4/BA.5-like immune escape variant, the previously observed protective effects of vaccination against the second wave were substantially reduced, as the efficacy against transmission and severe disease is impacted by the immune escape. For these immune escape scenarios, we observed a linear relationship between past attack rate and future attack rate. Furthermore, future attack rates are often *higher* than past attack rates (Fig. 5 a, b).

We found that older populations have slightly greater near-future attack rates given the same past hybrid immunity as younger populations, but overall we did not observe a substantial difference between peak sizes, similar to the previous scenarios where transmission is dominated by a single viral strain. For severe outcomes such as death in the second wave, we found that the protective effect of vaccination was substantially reduced, with much smaller differences in clinical outcomes between different vaccination coverages. Once again, older populations (i.e. populations with larger high-risk groups) experienced consistently worse clinical outcomes than the younger populations (Fig. 5 c, d).

## 4 Discussion

The Omicron variant of COVID-19 has spread across the world. Unlike previous variants, Omicron has increased transmissibility and immune escape [28, 29, 2, 30, 31], meaning that both vaccination and past infection confer a lower level of protection against Omicron (re)infection. We considered different populations with either ‘older’ or ‘younger’ demographics and with varying existing hybrid immunity. These populations were then subjected to a second wave of either the same prior variant (BA.1 like) or a different immune escape variant (BA.4/BA.5 like). We find that high vaccination coverage makes a noticeable difference in reducing the number of infections and severe outcomes in populations. Additionally, a population with a high past attack rate—reflecting a society with certain mixing and environmental factors that increase disease spread—is likely to have a high future attack rate and will benefit from higher vaccination coverage. In contrast, if populations with a low past attack rate—reflecting different intrinsic society mixing and limited disease spread, including potential public health measures—continue to behave the same going forward, then high vaccination does not have high impact against a second wave.

We find that higher vaccination coverage is more important for older populations, that is, populations with larger high-risk groups. Even if older and younger populations have the same future wave size—i.e. in terms of infection numbers—the older populations consistently have worse clinical outcomes. This work highlights that populations with large high-risk groups must do more to protect themselves in order to achieve the same outcomes compared to those populations with smaller high-risk groups.

We also found that high vaccination coverage can *delay* the emergence of a second wave. This delay is important because a longer time between waves allows more time to vaccinate populations, fewer infections and deaths across the same time frame, less economic disruption, and so forth. If there is a sufficient interval between campaigns, a fourth booster dose may be able to mitigate against some of these outcomes in the older population by “resetting” short term protection to a higher level, but that has not been simulated here. For example, Hogan et al. [34] created a model to predict how protection from past infection, vaccination and boosting declines over time. They found that in partially vaccinated populations, (first) boosters should be preferentially given to high-risk/older groups instead of giving new primary doses to low-risk/younger age groups; considering waning immunity, we can expect this reasoning to apply for second and subsequent boosters.

A strength of our study is that using our agent-based transmission model, we have been able to systematically model complex exposure histories for various populations with differing overall levels of waning hybrid immunity and compare how different populations fare under various scenarios. As vaccinations and exposures increase over time, the hybrid immunity landscape of populations will become more complex, further increasing the utility of models that have the ability to capture diverse exposure and vaccination histories. Many of the earlier studies did not include flexible hybrid immunity arrangements and/or did not include waning immunity or multiple reinfections (e.g. [44, 45, 43]), which here we show is key to the occurrence, size and timing of the second wave. While we did not include heterologous vaccination schedules, such as was modelled in [67], our modelling framework has the ability to support these scenarios.

There are several limitations to our study. First, our model did not have two variants circulating at the same time, instead using a sharp change point where all new infections following a certain time were with the immune escape strain. However, given that new variants have historically replaced currently circulating variants over relatively short time periods [68, 69], we do not envisage this would greatly change our results. Second, we did not consider the breadth of antibody response; we assume that while natural infection and vaccination boost antibodies by different amounts, the model then wanes both at the same rate instead of different rates. The next generation of vaccines will very likely be bivalent, as they are more broadly cross reactive and provide more durable protection. With all the increasing *breadth* of protection becoming important, a single antibody value may no longer be sufficient, instead requiring a multidimensional approach. Thirdly, in the absence of robust population level evidence to the contrary, our model assumes that neutralising antibody titres are the only proxy correlate of protection against both infection and clinical outcomes. However, we expect there exist other correlates of protection [70, 71], and as evidence regarding these mechanisms accrues, we will incorporate them into future model development.

Our study clearly shows how waning immunity and emergence of vaccine escape variants limits the impact of COVID-19 vaccines against transmission. The benefits of vaccination and past infection derived protection against COVID-19 disease depend critically on many factors, including demography, health systems capacity, vaccine efficacy, and breadth of protection, especially against new variants. Protection against severe disease is more robust, with achievable gains depend on underlying population demographics and risk. Ideally, vaccines would be allocated after assessing hybrid immunity at both the individual and population level in order to maximise the protection against future waves especially if vaccine stocks are limited. Our results suggest that populations with low vaccination coverage and high past infection rate should still consider vaccination if public health measures are not enforced or social mixing is not reduced.

The hybrid immunity landscape will only become increasingly complex as time goes on: with more circulating variants and with more vaccinations and more unique and individual exposure histories. How this can be robustly incorporated into mathematical models—especially in cases of limited available data—will be an ongoing challenge. As we move from the initial pandemic stage of COVID-19 to ongoing endemic transmission, we need tailored responses, sustainable long term protection and vaccination schedules to protect those at risk of severe disease and have equity of outcomes going forward. The role of future vaccines for resilience to ‘endemic’ disease and response to variants of concern remains to be determined for each individual country context.

## Data Availability

All data produced in the present study are available upon reasonable request to the authors. The code will eventually be available as a public repository.

## Acknowledgments

We thank Gerry Ryan for various neutralising antibody titre parameters used in the immunological model.

## Funding

This work was supported by the Australian Government Department of Foreign Affairs and Trade Indo-Pacific Centre for Health Security (Supporting Preparedness in the Asia-Pacific Region through Knowledge). ABH and IM are funded by an Australian National Health and Medical Research Council Investigator Grants.

## A Simulation details and results

### A.1 Constructing the exemplar ‘younger’ and ‘older’ populations

There are multiple methods to measure population aging, especially in light of increasing life expectancies and extended working age across the world [72, 73]. For our populations, we use the old-age to working-age demographic ratio, OADR, which is calculated as:

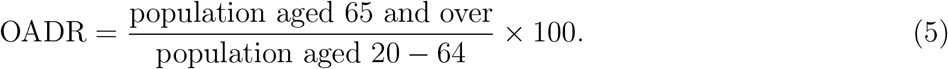

As mentioned in the main text, our exemplar “younger” and “older” populations are constructed from the averaged age distribution across multiple countries in the WHO-defined Western Pacific Regions. To obtain two different age distributions—which is also a proxy for size of high-risk group—we defined “younger” countries as having an *OADR* ≤ 12 and “older” countries as those with an *OADR* ≥ 15 (we do not include countries whose OADR lay between 12 and 15). Coupled with the international population data available at [55] for the year 2021, the countries we included in the “younger” and “older” groups were:

1. “Younger”: Mongolia, Brunei Darussalam, Cambodia, Lao People’s Democratic Republic, Philippines, Fiji, Papua New Guinea, Soloman Islands, Vanuatu, Kiribati, Micronesia (Fed. States of), Samoa, and Tonga (see Fig. 6).
2. “Older”: China, Hong Kong SAR, Macao SAR, Japan, Republic of Korea, Singapore, Australia, New Zealand, New Caledonia, Guam, and French Polynesia (see Fig. 7).

### A.2 Detailed vaccination allocation and schedule

There are two components to the vaccination program in our simulations: (1) the vaccine schedule, and (2) the vaccine allocation (which also defines the vaccine capacity).

The vaccine schedule has three different stages, each 26 weeks each—roughly half a year. An example of the schedule is given in Fig. 9.

1. In the first stage, first doses are given to the allocated groups that will be vaccinated during the first year. The 65+ individuals are vaccinated first, followed by randomly assigned vaccinations among the 5 *−* 64 age group.
2. In the second stage, the second dose is given to the same individuals as the first stage. The 65+ individuals receive their second doses first (though their exact dates are random). Once all the 65+ individuals that will be vaccinated are vaccinated, the 5 *−* 64 groups are vaccinated at random days. At the end of the second stage, *V* % of the population have received the primary course, where in the main paper, *V* = 20, 50, 80.
3. In the third stage, 80% of the fully vaccinated population receive a booster dose, while remaining available doses are given out as new primary course doses. During the third stage, 80% of daily doses are used as booster doses (for any age group 5+), while 20% of daily doses are used as new primary course doses (for any age group 5+).

**Figure 6:**
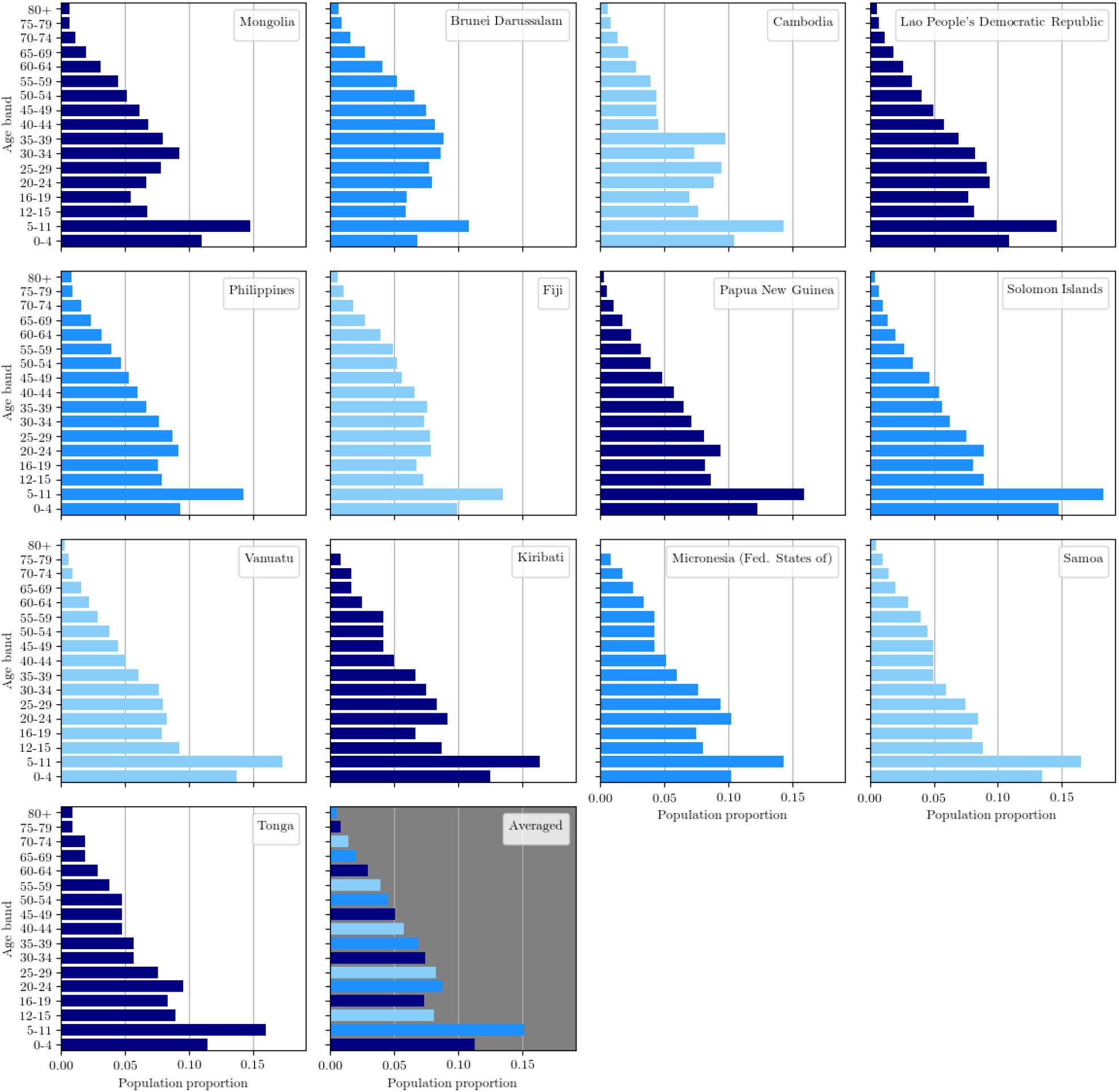
Countries used to create the exemplar ‘younger’ population distribution: relative proportions across age groups were averaged.

Suppose we want an initial *V* % vaccination coverage. We assume that the per-stage vaccination capacity is *V C* = 100, 000 *× V* % doses, that is, we will have *V C* doses per stage, or *V C/*26 doses per week. Thus, in the first stage, we have *V C* first doses. In the second stage, we have *V C* second doses. And in the third stage, we have *V C ×* 80% booster doses and *V C ×* 20% new primary course doses (which means *V C ×* 10% new first doses and *V C ×* 10% new second doses).

**Figure 7:**
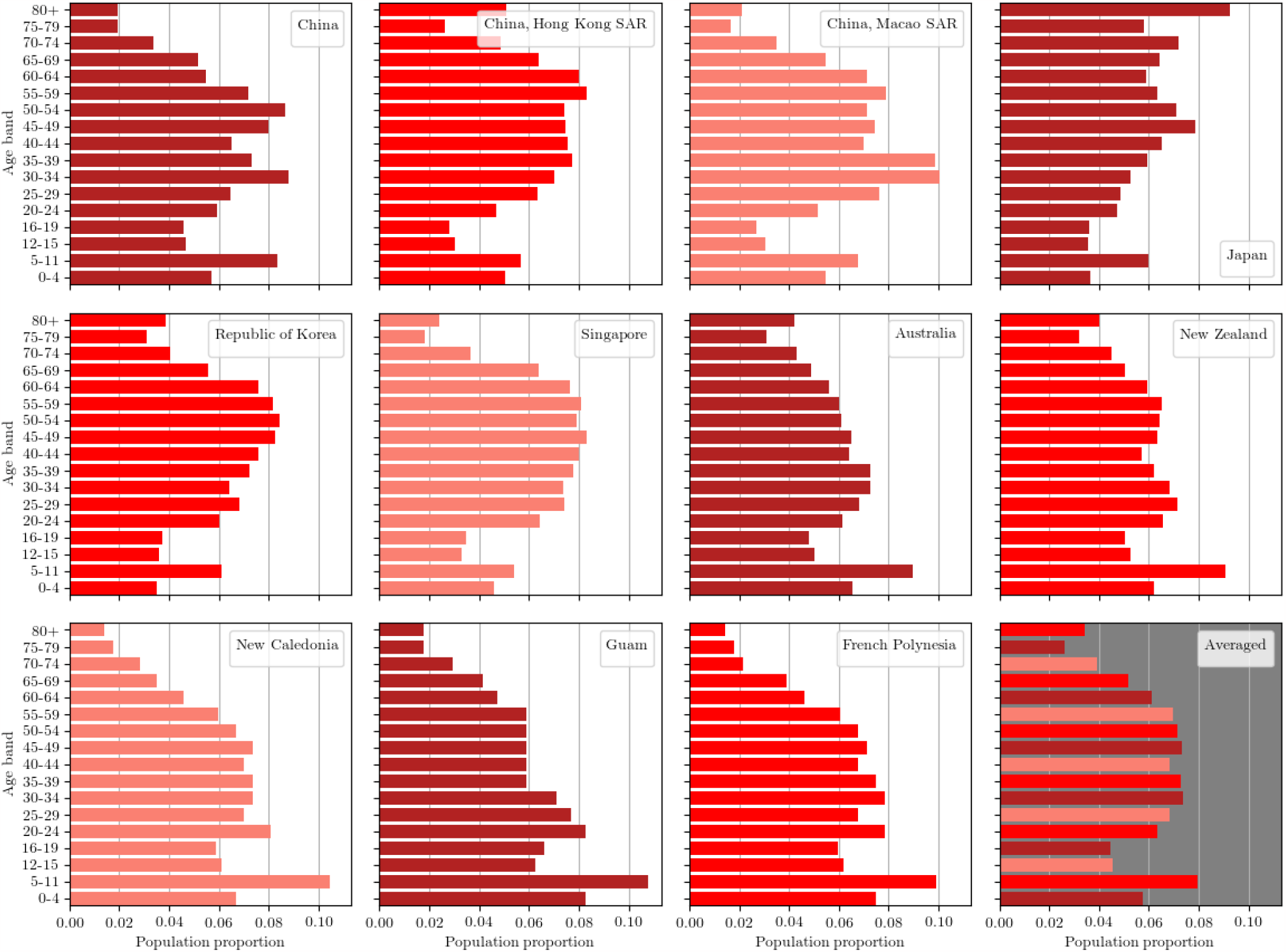
Countries used to create the exemplar ‘older’ population distribution: relative proportions across age groups were averaged.

The vaccine allocation broadly follows WHO guidelines, which recommends prioritising the vaccination of older and higher-risk groups. At the lower vaccination rate of 20%, we ensure 80% of the 65+ age group are fully vaccinated by the first year. At the higher vaccination rates of 50% and 80%, we ensure that 95% of the 65+ age group are fully vaccinated by the end of the first year. Remaining doses available are distributed evenly among the lower age groups. An example of this allocation is given in Fig. 10.

### A.3 Extended results

Fig. 11 shows the near-future ICU admissions for the various populations and scenarios. Overall, the trends in ICU admission reflect the trends for deaths: that older populations typically have worse outcomes, and that vaccination can modestly reduce ICU admission numbers in all scenarios provided that vaccination coverage was 50% or above.

**Figure 8:**
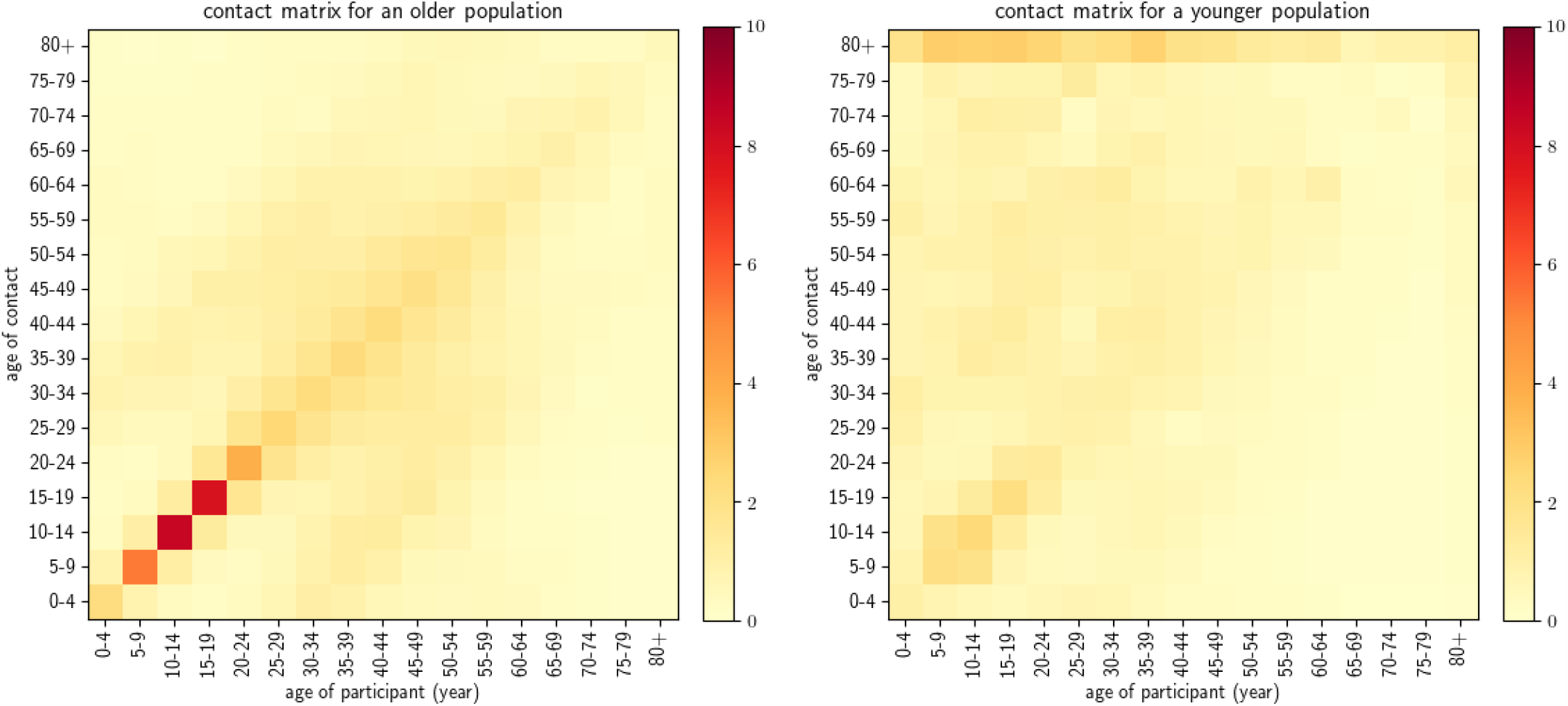
Contact matrices used.

Fig. 12 demonstrates the range of outcomes of multiple simulations. Fig. 12(a) and (b) show infections averted by vaccination compared with scenarios representing the baseline no-vaccination case, while Fig. 12(c) and (d) show deaths averted by vaccination compared with scenarios representing the baseline no-vaccination case.

At 80% vaccine coverage, the number of infections averted increases steadily in relation to the first wave attack rate which reflects the degree of transmission in the population. As vaccine coverage falls, the modelled maximum number of infections averted starts to decline for higher transmission pressures/attack rates.

Overall, in an older population more deaths can potentially be prevented by vaccination than in a younger population. At 80% vaccine coverage, the number of deaths averted increases steadily in relation to the first wave attack rate which reflects the degree of transmission in the population. As vaccine coverage falls, the modelled maximum number of deaths averted starts to decline for higher transmission pressures/attack rates.

Fig. 13 shows the avoided infections and avoided deaths due to vaccination compared with scenarios representing the baseline no-vaccination case, in the scenarios where the second wave is due to an immune escape variant. We see that as vaccination coverage increases, we have have increases in averted infections.

**Figure 9:**
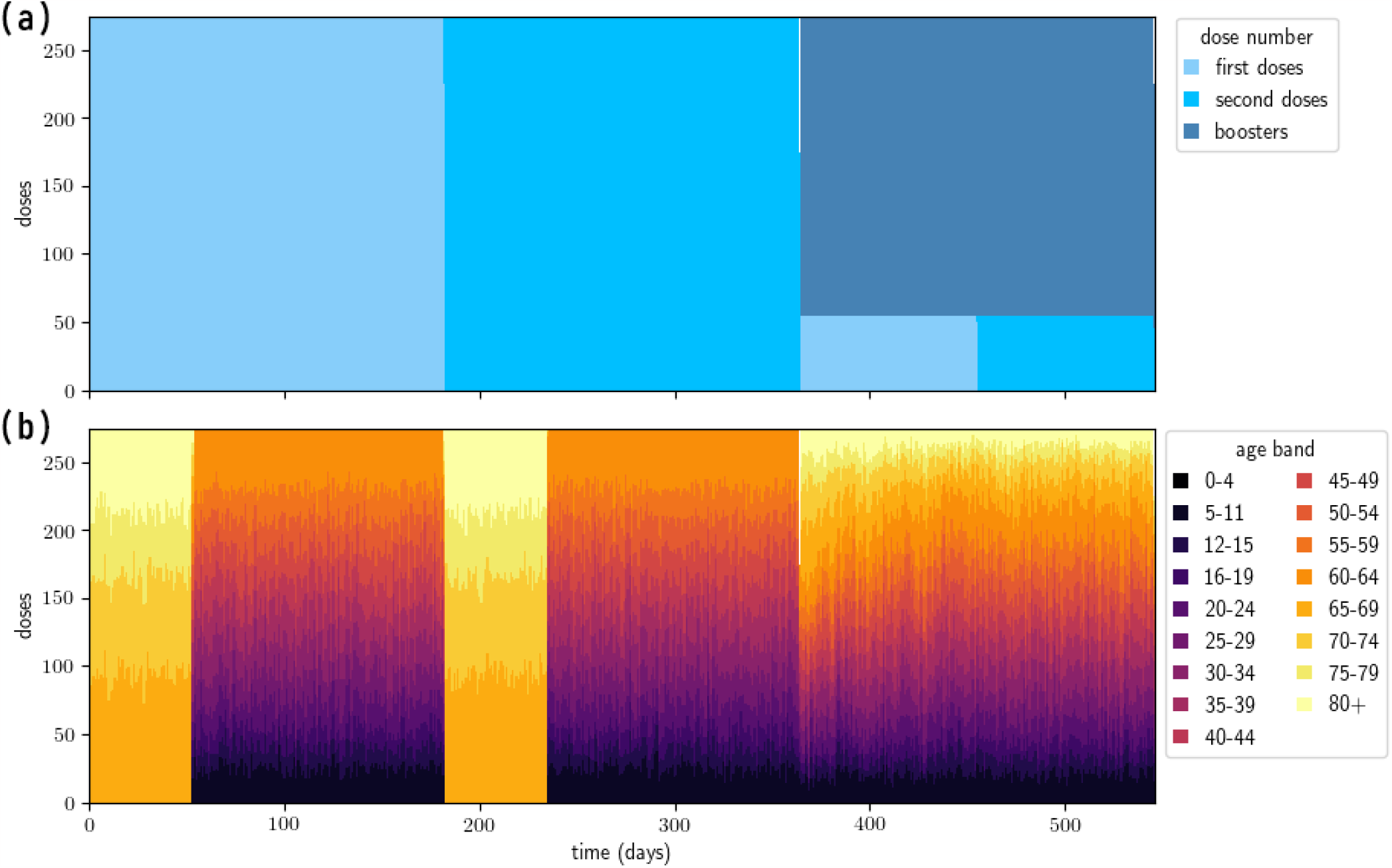
Vaccine schedule and distribution example: older population with 50% vaccination coverage after the first year. In the top panel, we see that doses are given out in the order of first doses, second doses, then jointly boosters and new primary doses. In the bottom panel, we see that during the first two stages, the older population 65+ are given their dose (numbered one or two) first, followed by the rest of the population. Meanwhile during the third (booster) stage, multiple different age groups are vaccinated daily, either with boosters or with new primary doses (with no age prioritisation for the booster dose).

### A.4 50% vs 80% booster coverage

In the main text, 80% of the initially vaccinated population were given boosters, while the remaining vaccination capacity was used to give new primary doses to previously unvaccinated individuals (note that all individuals vaccinated during the first year prior to boosters received the full primary course). In an alternative scenario, we could provide boosters to only 50% of the initially vaccinated population and try to vaccinate more people who had not yet been vaccinated. Fig. 14 shows the total difference in infection numbers and death numbers between the two scenarios, of which there is not a large noticeable difference.

**Figure 10:**
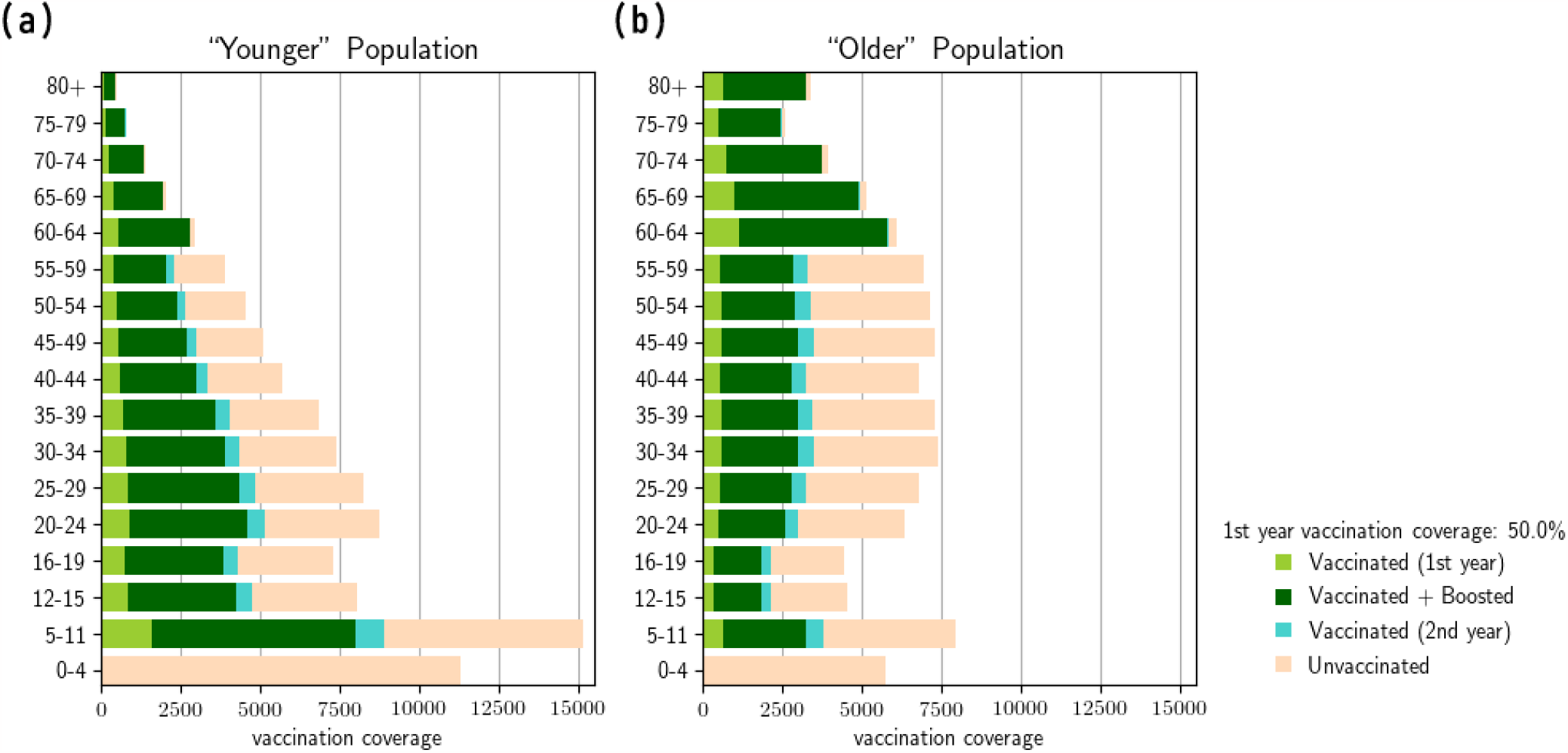
Vaccine distribution example: older and younger population with 50% vaccination coverage. The 65+ age group can be vaccinated to a maximum in-group coverage of 95%, while remaining available doses are distributed uniformly to the remaining 5-64 year old groups.

## B If vaccination efficacy was worse

Vaccine efficacy is important in determining just how much effect vaccination can have on future outcomes. In this section, we briefly consider a scenario where vaccine efficacy is lowered, with the new parameters given in Table 2. The results are given in Fig. 15. With a worse vaccine, we find less difference in future infection/future attack rate given different vaccine coverages. Compared to the results in the main paper with a better vaccine, in the scenarios here, there are higher future attack rates and also higher numbers of severe outcomes.

**Figure 11:**
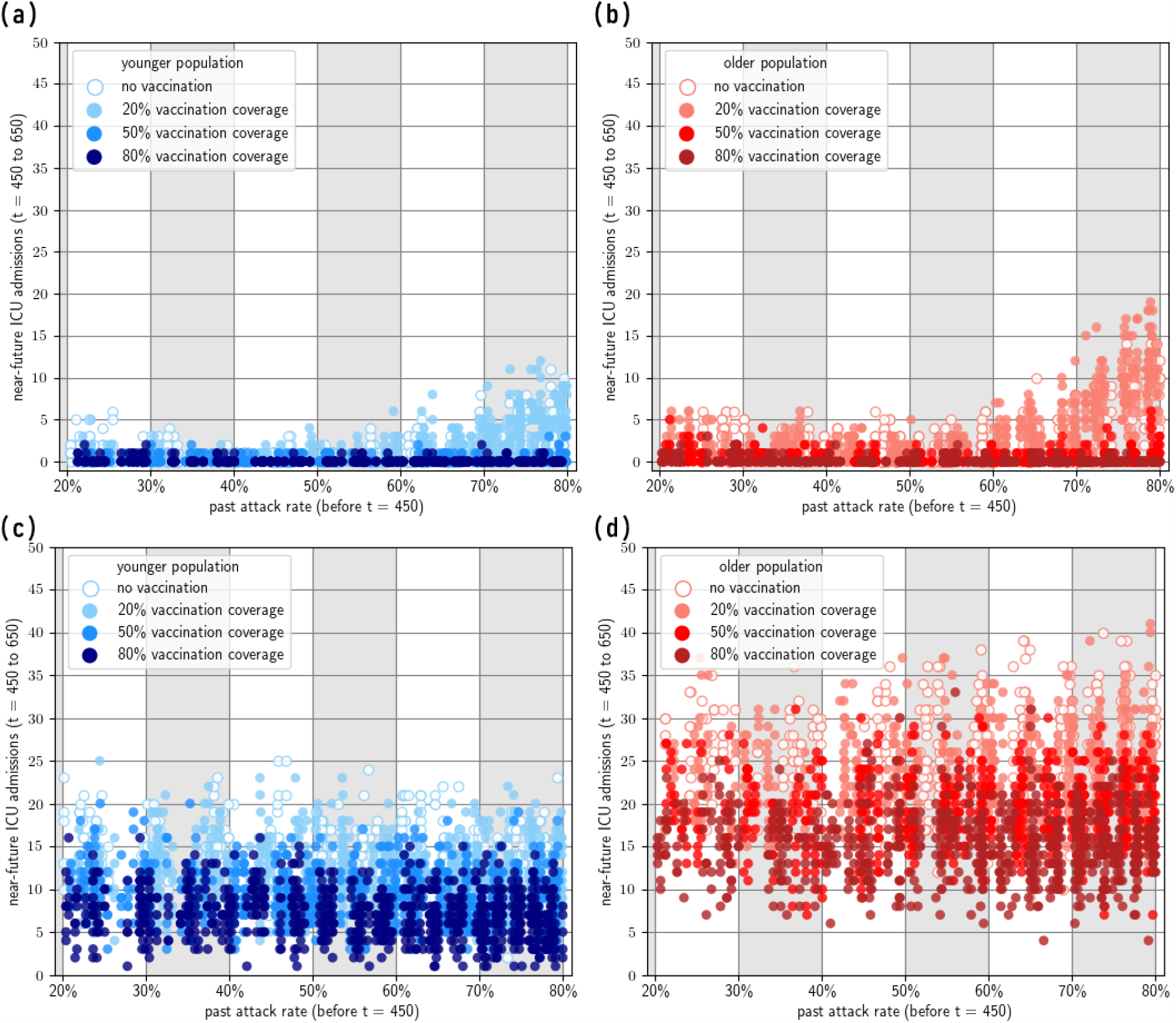
Near-future ICU admissions. (450 ≤ *t* ≤ 650) given past attack rate (*t <* 450). Increased vaccination coverage reduces the upper limit of ICU admissions. Top row: second wave due to the same variant. Bottom row: second wave due to a BA4/5-like variant.

**Figure 12:**
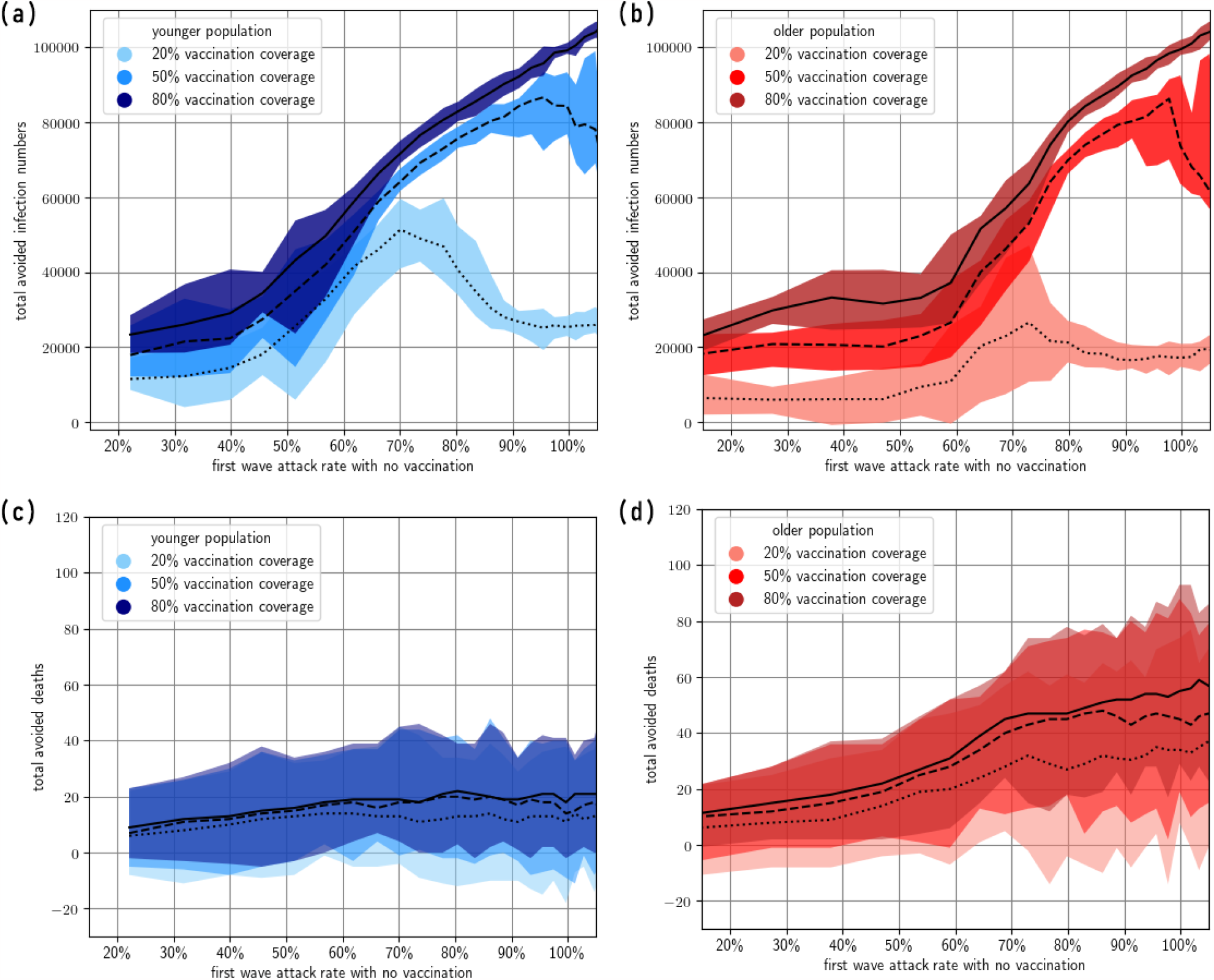
Total avoided infections and total avoided deaths (where the second wave is due to the same variant). We compare the vaccination scenarios with the scenario with no vaccination throughout the full simulation. The horizontal axis is the past (first wave) attack rate of the scenario with no vaccination. Note that the upper limit in total avoided deaths is the same within each plot (c) and (d), as there exists some simulations where no deaths occur. The width of each colour band reflects the minimum/maximum of simulations exploring three levels of vaccine coverage (the lines show the median).

**Figure 13:**
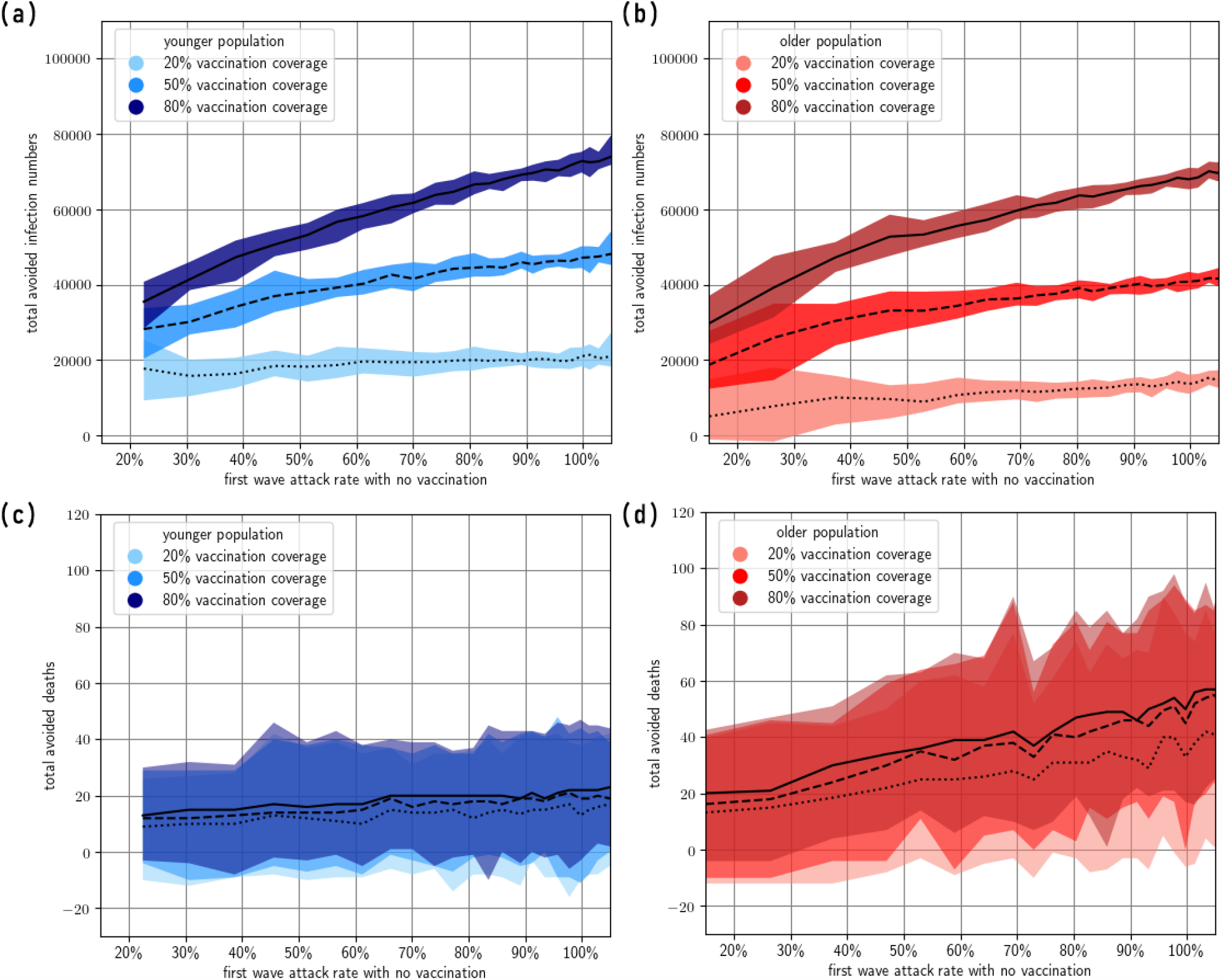
Total avoided infections and total avoided deaths (where the second wave is due to a BA4/5-like immune escape variant). We compare the vaccination scenarios with the scenario with no vaccination throughout the full simulation. The horizontal axis is the past (first wave) attack rate of the scenario with no vaccination. The width of each colour band reflects the minimum/maximum of simulations exploring three levels of vaccine coverage (the black lines show the median).

**Figure 14:**
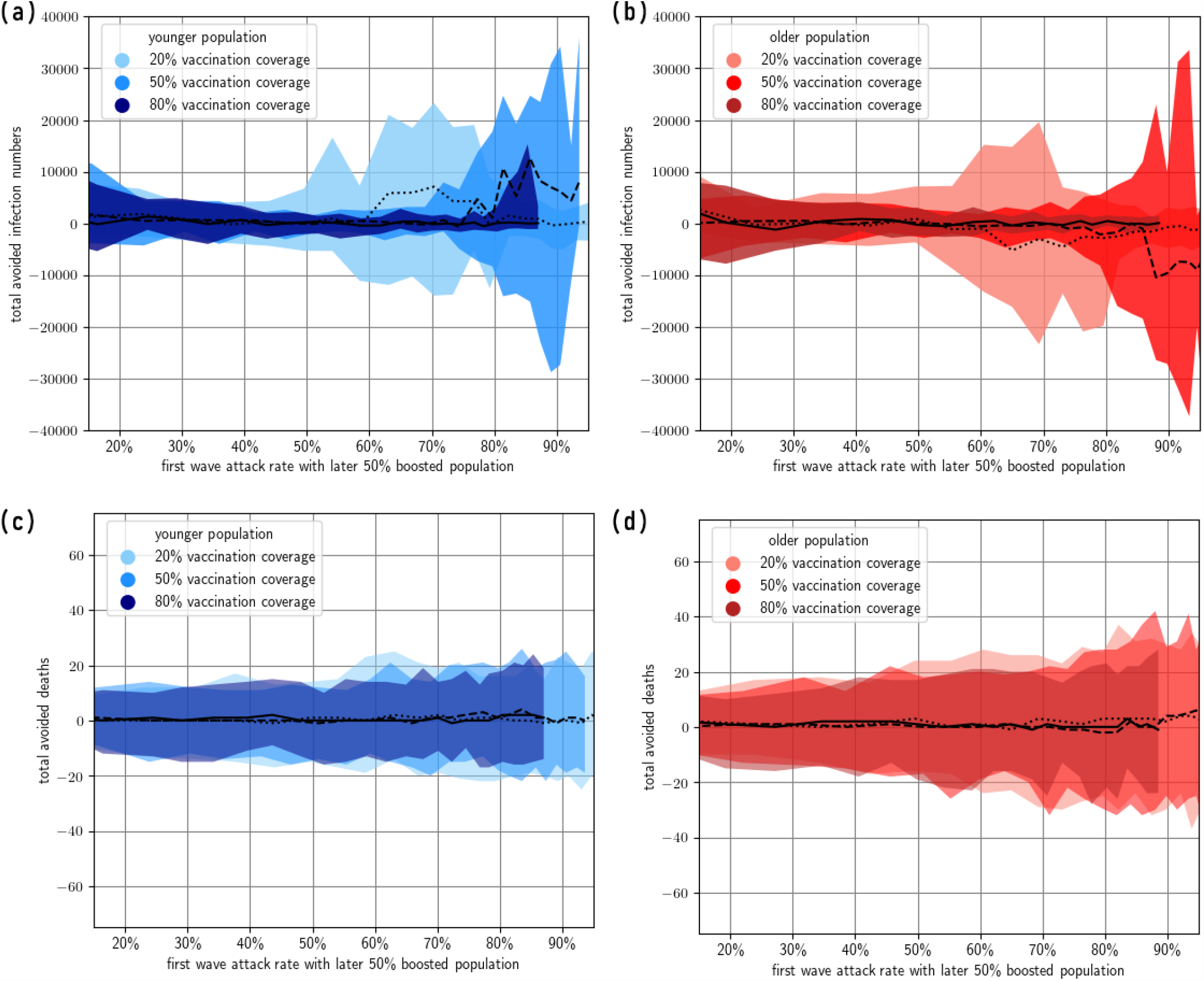
**Difference between infections and deaths for 50% vs 80% booster coverage allocation** (and where remaining doses are given out as new primary doses). Assumes that the second wave is due to the same variant.

**Table 2:** Changed model parameters used for a lower efficacy vaccine. Source: Ref. [53]

| Parameter: description | Value |
| --- | --- |
| $c_h$: $c_{50}$ of hospitalisation | -1.206211545 |
| $c_d$: $c_{50}$ of death | -1.183605217 |
| $c_\xi$: $c_{50}$ of acquisition | -0.471657741 |
| $c_\tau$: $c_{50}$ of transmission | 0.018459279 |
| $c_q$: $c_{50}$ of symptoms | -0.634901845 |
| $\mu_{AZ1}^0$: $\log_{10}$ of the mean neutralising antibody titre after the first dose of AstraZeneca | -1.318173846 |
| $\mu_{AZ2}^0$: $\log_{10}$ of the mean neutralising antibody titre after the second dose of AstraZeneca | -0.939262177 |
| $\mu_{P3}^0$: $\log_{10}$ of the mean neutralising antibody titre after the first booster dose of Pfizer | -0.336224683 |
| $\log(k)$: governs the steepness of the logistic curve relating antibodies to protection against disease outcome | 1.707366579 |
| $k_a$: decay rate of neutralising antibodies | 0.008502135 |
| $\mu_{\text{infection}}$: $\log_{10}$ of the mean neutralising antibody titre additionally gained after infection | 1.158362492 |

**Figure 15:**
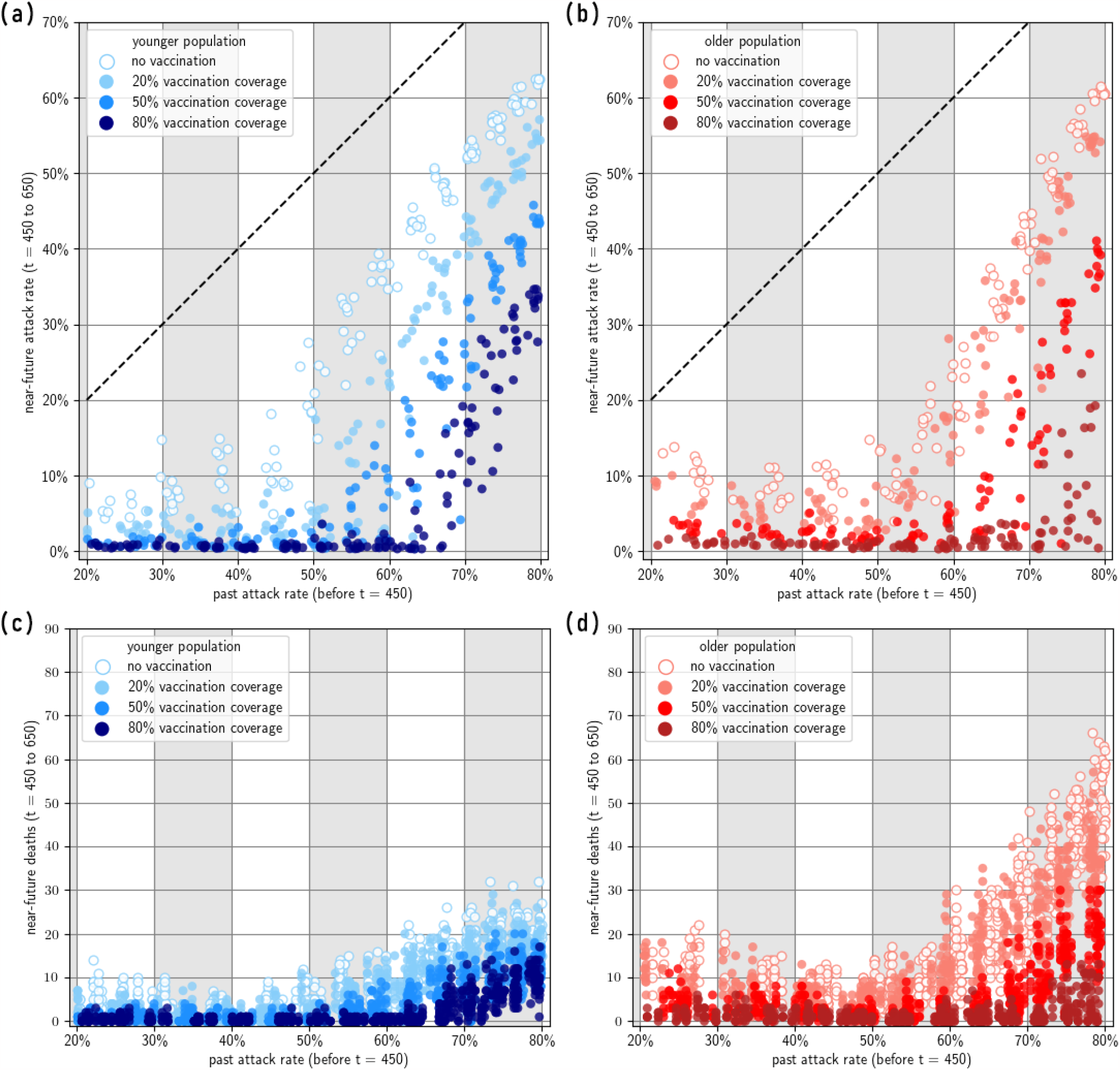
Near-future attack rate and deaths given past immunity, for a worse vaccine. Assuming a second wave due to the same Omicron BA.1 like variant.

## Notes

### Competing Interest Statement

The authors have declared no competing interest.

